# A Phase I Study of a PARP1-targeted Topical Fluorophore for the Detection of Oral Cancer

**DOI:** 10.1101/2020.11.09.20228536

**Authors:** Paula Demétrio de Souza França, Susanne Kossatz, Christian Brand, Daniella Karassawa Zanoni, Sheryl Roberts, Navjot Guru, Dauren Adilbay, Audrey Mauguen, Cristina Valero Mayor, Wolfgang A. Weber, Heiko Schöder, Ronald A. Ghossein, Ian Ganly, Snehal G. Patel, Thomas Reiner

## Abstract

**Purpose:** Visual inspection and biopsy is the current standard of care for oral cancer diagnosis, but is subject to misinterpretation and consequently to misdiagnosis. Topically applied PARPi-FL is a molecularly specific, fluorescent contrast-based approach that may fulfil the unmet need for a simple, *in vivo*, non-invasive, cost-effective, point-of-care method for the early diagnosis of oral cancer. Here, we present results from a phase I safety and feasibility study on fluorescent, topically applied PARPi-FL.

**Patients and Methods:** Twelve patients with a histologically proven squamous cell carcinoma of the oral cavity (OSCC) gargled a PARPi-FL solution for 60 seconds (15 mL, 100 nM, 250 nM, 500 nM, or 1000 nM), followed by gargling a clearing solution for 60 seconds. Fluorescence measurements of the lesion and surrounding oral mucosa were taken before PARPi-FL application, after PARPi-FL application and after clearing. Blood pressure, oxygen levels, clinical chemistry and CBC were obtained before and after tracer administration.

**Results:** PARPi-FL was well-tolerated by all patients without any safety concerns. When analyzing the fluorescence signal, all malignant lesions showed a significant differential in contrast after administration of PARPi-FL, with the highest increase occurring at the highest dose level (1000 nM), where all patients had a tumor-to-margin fluorescence signal ratio of > 3. A clearing step was essential to increase signal specificity, as it clears unbound PARPi-FL trapped in normal anatomical structures. PARPi-FL tumor cell specificity was confirmed by *ex vivo* tabletop confocal microscopy. We have demonstrated that the fluorescence signal arose from the nuclei of tumor cells, endorsing our macroscopic findings.

**Conclusions:** A PARPi-FL swish & spit solution is a rapid and non-invasive diagnostic tool that preferentially localizes fluorescent contrast to OSCC. This technique holds promise for the early detection of OSCC based on *in vivo* optical evaluation and targeted biopsy of suspicious lesions in the oral cavity.

**Translational Relevance:** Despite their accessible location, oral cavity cancers are often diagnosed late, especially in low-resource areas where their incidence is typically high. The high prevalence of premalignant and benign oral lesions in these populations contributes to a number of issues that make early detection of oral cancer difficult: even in experienced hands, it can be difficult to differentiate cancer from premalignant or benign lesions during routine clinical examination; and biopsy-based histopathology, the current standard of care, is invasive, prone to sampling error, and requires geographic access to appropriate health care professionals, including a highly trained pathologist. While seemingly impenetrable economic and infrastructure barriers have confounded the early diagnosis of oral cancer for most of the world’s population, these could be circumvented by a simple, *in vivo*, non-invasive, cost-effective, point-of-care method of diagnosis. We are attempting to address this unmet clinical need by using topically applied PARPi-FL — a molecularly specific, fluorescent contrast-based approach — to detect oral cancer.

**Funding:** This work was supported by National Institutes of Health grants P30 CA008748, R01 CA204441 (TR) and R43 CA228815 (CB and TR). Dr. Valero was sponsored by a grant from Fundación Alfonso Martín Escudero. The funding sources were not involved in study design, data collection and analysis, writing of the report, or the decision to submit this article for publication.

**Disclosure of Potential Conflicts of Interest:** C.B., S.K., S.P. and T.R. are shareholders of Summit Biomedical Imaging, LLC. S.K., S.P. and T.R. are co-inventors on PCT application WO2016164771. T.R. is co-inventor on PCT application WO2012074840. T.R. is a paid consultant for Theragnostics, Inc. All the other authors have no relevant conflict to declare. This arrangement has been reviewed and approved by Memorial Sloan Kettering Cancer Center in accordance with its conflict of interest policies.

## Introduction

Worldwide, oral cancer is the 7^th^ most frequent cancer and is responsible for approximately 355,000 newly diagnosed cases and 177,000 deaths per year (1). Squamous cell carcinoma of the oral cavity (OSCC), accounts for over 90% of all cases (2). Although the oral cavity is an anatomical site that is easily accessible for examination by both patients and health care providers, only 29% of patients in the US are diagnosed when the tumor is still localized. Early diagnosis is associated with a 5-year survival of 85%. With advancing stages, survival rates drop steeply to 48% with regional involvement, and 19% when distant metastasis are present (3). In addition to poorer survival results, patients with more advanced cancers also suffer from poorer quality of life after treatment because they need more extensive surgery and postoperative adjuvant treatment. Early diagnosis is therefore an essential determinant both of better survival and post-treatment quality of life in patients with OSCC.

The current standard of care for evaluation of oral lesions is visual inspection followed by histopathological examination of a biopsy from the lesion based on the level of suspicion for malignancy. There are a number of barriers to early diagnosis of OSCC, including those related to demographic and patient factors (4), physician factors (5), and limitations of currently available diagnostic tests (6).

Despite their accessible location, oral cavity cancers are often diagnosed late, especially in low-resource areas where their incidence is typically high. The high prevalence of premalignant and benign oral lesions in these populations contributes to a number of issues that make early detection of oral cancer difficult: even in experienced hands, it can be difficult to differentiate cancer from premalignant or benign lesions during routine clinical examination; and biopsy-based histopathology, the current standard of care, is invasive, prone to sampling error, and requires geographic access to appropriate health care professionals, including a highly trained pathologist. Premalignant lesions are often misinterpreted on visual examination as benign mouth ulcers or inflammation, delaying their correct diagnosis (7). Another complicating factor is that premalignant lesions, for example erythroplakia and leukoplakia, do not have uniform rates of progression to cancer (ranging from 6%–36%) and are often accompanied by varying grades of dysplasia (mild/moderate/severe) (8). The lack of a precise diagnostic test and reliable estimation of cancer risk poses challenges in designing effective strategies for follow-up of suspicious lesions and for early diagnosis of OSCC.

Tissue biopsy followed by histopathological assessment remains the current diagnostic standard. While biopsies yield a reliable diagnosis, their findings represent only a single assessment in space and time, and allow neither extrapolation to the entire lesion due to sampling limitations nor follow-up without additional biopsies. This is especially important for patients who harbor multiple lesions, and for whom pathologic assessments of every lesion via biopsy would be impractical.

Our group and others have reviewed the extensive available literature on preclinical and clinical studies that investigate current methods for oral cancer detection (2,9,10). These include cytology-based evaluation methods (e.g. brush biopsy (11)), staining methods with acetic acid (12), toluidine blue (13), Lugol’s iodine (12), and light-based detection systems, including reflectance confocal microscopy (14), chemiluminescence (11) and autofluorescence (15). However, due to many impenetrable economic and infrastructure barriers, none of these techniques have become established as reliable tools for screening and guiding treatment decisions for most of the world’s population. This underlines the need for simple, *in vivo*, non-invasive, cost-effective, point-of-care intraoral imaging agents that have high specificity and sensitivity and low false positive rates, which are therefore able to aid in screening and early detection of OSCC.

In this Phase I, dose escalation clinical trial, we have explored the use of a fluorescent imaging agent, PARPi-FL (16-20), as a tool for non-invasive *in vivo* identification of malignant oral lesions in real time (**Fig. 1A**). PARPi-FL targets poly-ADP ribose polymerase 1 (PARP1), a key enzyme in the DNA damage response, which is overexpressed in tumors due to increased proliferation, mutational burden and genomic instability (21-24). PARP1 overexpression occurs in many tumors, including in malignant oral lesions compared to normal mucosa and dysplastic lesions (20,25). We were able to show that with PARPi-FL, tumor could be distinguished from normal tissue in human oral cancer specimen with sensitivity and specificity of greater than 95%, using an PARPi-FL staining approach that does not require freezing or fixation of tissue (25). PARPi-FL is particularly interesting for oral cancer screening, because it is cell and tissue permeable, does not intercalate DNA and can be applied topically, for example via gargling, penetrating tumor tissue within minutes (18,25,26) (**Fig. 1B**).

**Figure 1.**
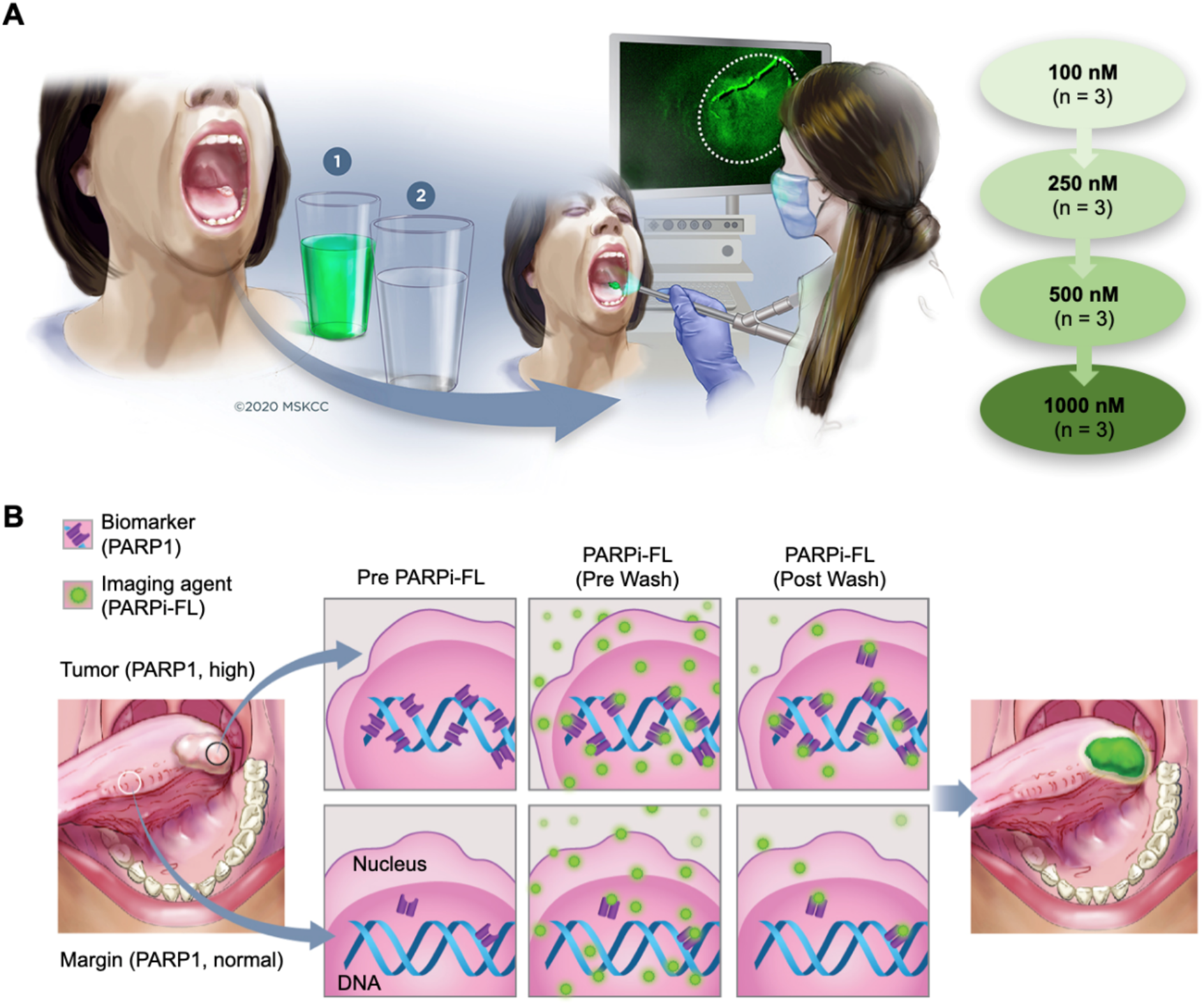
Design of phase I dose escalating study NCT03085147 and concept of PARPi-FL imaging for delineation of oral cancer. **A**, Patients (n = 12) with biopsy proven oral squamous cell carcinoma (OSCC) gargled a solution of PARPi-FL (step 1) at increasing concentrations (100 nM, 250 nM, 500 nM, and 1000 nM; each dose n = 3) for 1 min. Then, patients gargled a clearing solution for 1 min (step 2). Using a Quest Spectrum imaging device with an endoscopic camera and PARPi-FL optimized LED–filter system, the tumor area and surrounding margin of the patient were imaged before PARPi-FL administration, after PARPi-FL administration pre-wash and after PARPi-FL administration post-wash. **B**, The illustration describes the mechanism-of-action of PARPi-FL based oral cancer detection. After topical application of PARPi-FL in the oral cavity, the imaging agent binds to PARP1 in the nucleus of a cell. PARP1 is overexpressed in OSCC compared to normal mucosal tissue.

In the present study, patients with confirmed OSCC were imaged with a custom-made fluorescence equipped endoscope after topical application of PARPi-FL. We evaluated the safety of PARPi-FL topical application and analyzed the fluorescence signal in tumor and surrounding normal tissue by calculating tumor to healthy mucosal contrast ratios and assessing the feasibility of PARPi-FL imaging for oral cancer detection. In addition, we report initial results from a PARPi-FL phase II study, which evaluates the specificity of PARPi-FL for PARP1 in tumor cells.

### Patients and Methods

#### Study design

This exploratory, phase I, single-center, open-label, prospective Health Insurance Portability and Accountability Act compliant study was approved by the MSK Institutional Review Board and conducted in accordance with the Declaration of Helsinki (Clinicaltrials.gov - NCT03085147). Written informed consent was obtained from all patients. Patients were ≥ 18 years old, had a histologically or cytologically proven squamous cell carcinoma of the oral cavity (OSCC) and were scheduled to undergo surgery at MSK. Patients with any TNM (8^th^ edition) stage tumor were eligible for inclusion but needed to have an ECOG performance status of 0 or 1. Exclusion criteria were previous definitive surgical therapy (but not surgical biopsy) to the oral cavity or pharynx within the 2 weeks prior to PARPi-FL imaging, patients that underwent prior or ongoing treatment with a PARP1 inhibitor or presented a known hypersensitivity to Olaparib or to PEG 300. The primary objectives of this Phase I trial were to evaluate the safety and feasibility of PARPi-FL imaging. We also report the PARPi-FL-related fluorescence signal in the tumor compared to the normal surrounding tissue before and after topical exposure to PARPi-FL. Twelve patients were enrolled on this study protocol, 3 for each concentration level of the dose escalation (100 nM, 250 nM, 500 nM, and 1000 nM). All patients completed the study. The study design is summarized in **Fig. 1A**, and study procedures are summarized in **Fig. 2**. We have also imaged 1 patient from an ongoing PARPi-FL phase II study. This patient was imaged in the operating room, after administration of 1000 nM of PARPi-FL and subsequent clearing with 30% PEG 300 in water. A biopsy was taken from the tumor and from the free-of-disease margin for fluorescence microscopy and histopathological correlation.

**Figure 2.**
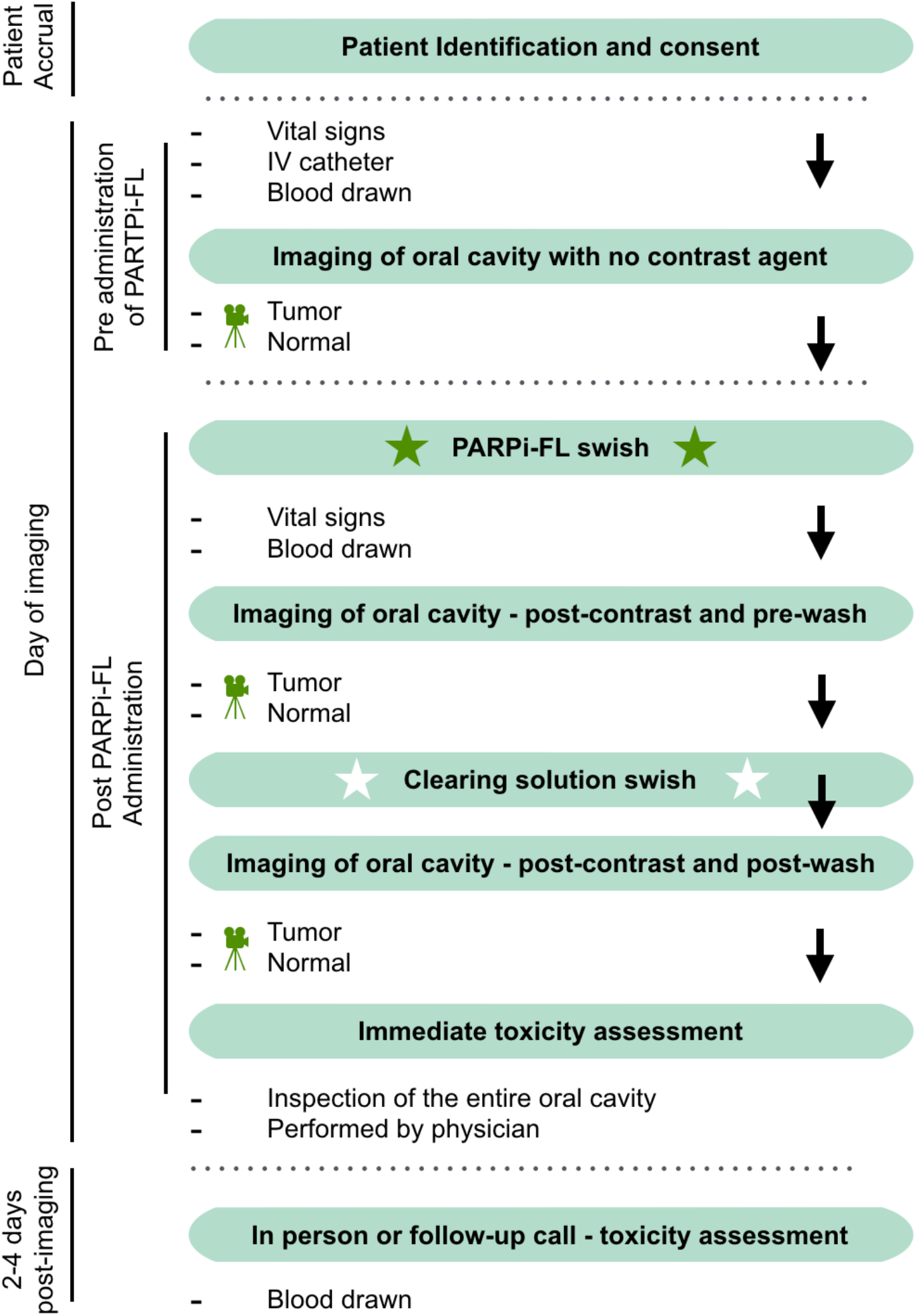
Schematic overview and flow chart of the PARPi-FL clinical trial. Twelve Patients with biopsy proven OSCC were identified and consented to the Phase I dose escalation study.

### Procedures

All patients underwent clinical examination and collection of vital signs (heart and respiratory rate, body temperature and blood pressure) before and after PARPi-FL administration. Intravenous blood samples (3 mL) were collected for hematology and biochemistry prior to PARPi-FL exposure and 2 - 4 days after the procedure (“clinical blood sample 1 and 2”). Another blood sample was collected between PARPi-FL exposure and imaging to evaluate whether PARPi-FL was present in a patient’s plasma. For PARPi-FL application (“swish”), the patient gargled 15 mL of the PARPi-FL containing solution for 60 seconds, and then spit it out. For the clearing step, the patient gargled 15 mL of the clearing solution for another 60 seconds and spat it out. The first 3 patients in the protocol (25% of the cohort) gargled a solution of 1% acetic acid in water. Due to taste complaints, the protocol was amended and the following 9 patients (75% of the cohort) gargled a solution of 30% PEG 300 in water. For the patient on phase II, a fresh tissue biopsy was taken from the tumor and from the free-of-disease margin and both were stained with a solution of 10% Hoechst 33342 in PBS before evaluation with confocal microscopy.

### Toxicity assessment

Vital signs were obtained before PARPi-FL administration and after completion of the imaging study. The patient’s oral mucosa was examined by the patient’s surgeon before PARPi-FL administration and immediately after completion of imaging for local irritation. A clinical exam or follow-up phone interview (up to 3 days after the imaging study) was carried out to document any possible side effects occurring after completion of the imaging study. Complete blood count and differential, routine clinical chemistry and a metabolic panel was obtained from the clinical blood samples.

### Chemicals and materials used for synthesis and vialing of PARPi-FL

Triethylamine (NEt_3_) >99.5% purity, Trifluoroacetic acid (TFA) ReagentPlus 99%, and dimethyl sulfoxide-d_6_ (DMSO-d_6_) were purchased from Sigma-Aldrich (St. Louis, MO). Anhydrous acetonitrile (CH_3_CN) over molecular sieve, HPLC-grade acetonitrile, BODIPY FL NHS ester, and Texwipe Technicloth were purchased from Thermo Fisher Scientific (Waltham, MA). Water (18.2 MΩ cm^-1^ at 25 °C) was obtained from an Alpha-Q Ultrapure water system from Millipore (Bedford, MA). PARP-NH precursor (4-(4-fluoro-3-(piperazine-1-carbonyl)benzyl)phthalazin-1(2H)-one) was synthesized at MSK according to procedures reported earlier (16,27).

### GMP synthesis of bulk PARPi-FL

PARPi-FL was produced under non-sterile good manufacturing practice (GMP) conditions at MSK under IND number 133,109. Briefly, BODIPY-FL NHS ester (5.0 mg, 12.8 μmol, 1.0 eq.) was conjugated to 4-(4-fluoro-3-(piperazine-1-carbonyl)benzyl) phthalazin-1(2H)-one (9.4 mg, 25.6 μmol, 2.0 eq.) in the presence of Et_3_N (6.3 μL) in anhydrous acetonitrile for 4 h at room temperature. Purification by preparative HPLC (Phenomenex Jupiter 5u C18 300A, 10 × 250 mm, 3 mL/min, 5 to 95% of acetonitrile (0.1% TFA) in 15 min) and subsequent lyophilization yielded PARPi-FL (5.9 mg, 73%) as a red solid. Analytical HPLC analysis (Waters’ Atlantis T3 C18 5 μm, 4.6 × 250 mm, 3 mL/min, 5 to 95% of acetonitrile (0.1% TFA) in 15 min) showed high purity (99.27%, *t*_*R*_ = 14.2 min) of the imaging agent. The identity of PARPi-FL was confirmed using LC-MS (MS(+) *m/z* = 621.15 [M-F]^+^) and NMR (consistent with structure). For reconstitution, polyethylene glycol (PEG) 300 (4.5 mL) was added to the lyophilized PARPi-FL followed by the addition of sterile saline (10.5 mL). The resulting PARPi-FL swish & spit solution (15 mL) was provided to the clinical research team and used within 12 hours after reconstitution. For the clearing solution, a 1% acetic acid solution Microlux/DL from Addent (Danbury, CT) was used in the first 3 patients. Then, the 1% acetic acid solution was replaced by a solution of 30% PEG 300 in water (15 mL) for the remaining 9 patients.

### Fluorescence imaging system

For PARPi-FL imaging, we used a Quest Spectrum imaging system (Quest Medical Imaging, Middenmeer, Netherlands) with a customized fluorescein isothiocyanate (FITC) light emitting diode (LED) attached to a 30° rigid laryngoscope was used. The collected images were post-processed to separate specific (PARPi-FL, sensor A) and non-specific (autofluorescence, sensor B) emissions based on different peak widths of the emission spectra (Supplementary Fig. 1). Only PARPi-FL related signals were further analyzed.

### Optimization of fluorescence imaging

Videos of agarose phantoms containing PARPi-FL (5.0 μM, 2.5 μM, 1.25 μM, 0.63 μM, 0.32 μM, 0.16 μM, and 0 μM) were acquired using the customized fluorescence endooscope in order to determine sensitivity and suitable imaging settings for the clinical study. Different frame rates (12 ms, 30 ms, 50 ms, and 83 ms) and distances from camera to phantoms (23 mm and 5 mm) were compared. Laser power was set to 100% and gain to 25.5 dB throughout the experiments. Still frames were selected from the videos and fluorescence intensity was quantified in 3 regions of interest using Fiji (ImageJ).

### Fluorescence imaging and analysis in patients

Videos were acquired before and at 2 time points post PARPi-FL application (pre-wash and post-wash) using the fluorescence laryngoscope. The same instrument settings were used for all imaging procedures (30 ms exposure time, 100% excitation power, gain: 25.5 dB). Furthermore, distance to the tissue surface was kept at around 10 mm. The videos were acquired by slowly scanning the tumor area and surrounding normal mucosa (non-tumor region). During this process, the real-time white light image was used for visual guidance and focusing. For image analysis, Quest QIFS Manipulation tool software (Quest Medical Imaging, Middenmeer, Netherlands) was used. We applied the autofluorescence subtraction tool to all patient images. For quantification, 3 still frames were selected from each video. In each frame, 5 regions of interest (ROI) were placed on tumor and non-tumor regions using Fiji (ImageJ). The white light channel images were used to determine ROIs of tumor and of normal mucosa (non-tumor regions). ROIs were then transferred to the fluorescence channel for PARPi-FL quantification. Tumor to non-tumor ratio was calculated by dividing each tumor ROI by the average of non-tumor ROIs in its respective frame.

### Analysis of PARPi-FL in blood

PARPi-FL content was analyzed in the research blood samples. To extract PARPi-FL, 4.0 mL of blood was added to 4.0 mL of a 1:1 ice-cold solution of acetonitrile and dimethyl sulfoxide (DMSO). The solution was vortexed for 30 seconds and centrifuged at 4,000 rpm for 10 min at 4 °C. The supernatant was isolated and ice-cold CH_3_CN (2.0 mL) was added. The solution was vortexed for 30 seconds and centrifuged at 4,000 rpm for 10 min at 4 °C. The supernatant was isolated and stored at −80 °C. After lyophilization and reconstitution in acetonitrile (200 μL), the solution was analyzed by Liquid Chromatography Mass Spectrometry (LCMS) for PARPi-FL detection (Supplementary Fig. 2).

### Pathology assessment

All patients underwent standard-of-care surgery after PARPi-FL imaging and the surgical specimens were processed as per routine protocol by a Head and Neck pathologist.

### PARP1 immunohistochemistry

Paraffin embedded sections of tumor and non-tumor research biopsies were available from 8 patients. PARP1 immunohistochemistry (IHC) and PARP1 quantification were performed according to our previously described procedure (20). We used an anti-PARP1 rabbit monoclonal antibody (46D11, Cell Signaling Technology, Danvers, MA) at 0.4 μg/mL, followed by a biotinylated goat anti-rabbit IgG (PK6106, Vector Labs, Burlingame, CA) at a 1:200 dilution. Hematoxylin & Eosin (H&E) stained slides from the same patients were used to determine areas of tumor, normal mucosal epithelium and normal muscle. Those exact areas were then referenced on a consecutive slide and were used for PARP1 quantification.

### Confocal Imaging

A fresh biopsy of the tumor and from adjacent non-tumor region was available in one patient in the phase II study (administered 1000 nM solution of PARPi-FL and the clearing solution). These tissues were further stained with a solution of 10% Hoechst 33342 in phosphate-buffered saline (PBS) and placed over a cover slip glass slide (48 × 60 mm no.1 thickness, Brain Research Laboratories, Newton, MA). Confocal images were acquired using a scanning confocal microscope (LSM880-Live, Zeiss, Germany) - 488 nm laser excitation for PARPi-FL (green), 405 nm for Hoechst (blue) and 561 nm (red) for autofluorescence. Images were further analyzed using our previously described Fiji (ImageJ) automatic script (28,29). Quantification of intensity of PARPi-FL signal was calculated by placing the region of interest on the Hoechst 33342 nuclear stain and calculating the signal corresponding to the same area using the green channel.

### Statistical analysis

Statistical analysis was carried out using R v3.6.0 and GraphPad Prism 8. Linear regression was used for PARP1 quantification on IHC and for the PARPi-FL inter-patient analysis. We used the Wilcoxon test (analysis of paired samples) to analyze vital signs and PARPi-FL imaging of intra-patient differences. Bar graphs display mean values, standard deviations and data points. Results with p-values < 0.05 were considered statistically significant and the level of significance for each result are displayed as *p < 0.05, **p < 0.01, ***p < 0.001, and ****p < 0.0001.

## Results

### Patient distribution

A total of 13 patients were included in this analysis. In the phase I part of the study, 12 patients were enrolled over a 29 months period (from November of 2017 to March of 2020). All patient data are summarized in **Table 1**. The median age was 65 years old (range: 48 - 80 years old) and 58% were male (n = 7). Seven patients (58%) were never smokers, whereas the rest had discontinued smoking. Seven patients were currently consuming alcohol (one of them only sporadically and the other 6 daily). Three patients had discontinued the use of alcohol and two reported that they never consumed it. All patients had a primary oral tongue tumor as the oral cavity subsite (n = 11 on the lateral tongue and n = 1 on the midline of the dorsal tongue) with a median size of 2.1 cm (range: 0.5 – 4 cm). Fifty percent (n = 6) of the lesions were exophytic, 25% (n = 3) had a flat morphology, 17% (n = 2) were ulcerative and one patient (8%) presented with a submucosal tumor. Patient 5 had an excisional biopsy of a squamous cell carcinoma of the lateral tongue at an outside hospital prior to referral to MSK. This patient was imaged with PARPi-FL and had a partial glossectomy performed 4 days later. The final pathological report of this patient revealed that there was no residual tumor in the glossectomy specimen. The one patient included in this report from the phase II study was enrolled in October of 2020.

**Table 1.** Patient distribution. Age, social habits, lesion location and T staging of patients enrolled in this study (n= 12, TNM 8^th^ edition).

| **Phase I - Patient distribution** |  |  |  |  |  |  |  |
| --- | --- | --- | --- | --- | --- | --- | --- |
| Case | Conc.<br>(nM) | Tabaco<br>Use | Alcohol<br>Use | Tongue<br>Sub-site | Lesion<br>Characteristic | Size<br>(cm) | Staging<br>(8<sup>th</sup> ed.) |
| 1 | 100 | never | current | lateral | exophytic | 3.0 | 2 |
| 2 | 100 | discontinued | discontinued | lateral | exophytic | 4.0 | 2 |
| 3 | 100 | never | current | lateral | ulcer | 3.0 | 2 |
| 4 | 250 | never | current | lateral | exophytic | 2.0 | 1 |
| 5 | 250 | discontinued | current | lateral | flat leukoplakia* | 1.0 | 1 |
| 6 | 250 | discontinued | never | lateral | exophytic | 1.5 | 1 |
| 7 | 500 | discontinued | discontinued | lateral | exophytic | 3.0 | 2 |
| 8 | 500 | never | current | lateral | ulcer | 2.3 | 2 |
| 9 | 500 | discontinued | discontinued | middle | submucosal | 1.1 | 1 |
| 10 | 1000 | never | never | lateral | exophytic | 0.5 | 1 |
| 11 | 1000 | never | occasionally | lateral | flat leukoplakia | 1.6 | 1 |
| 12 | 1000 | never | current | lateral | flat leukoplakia | 2.2 | 2 |

\*No tumor was found on the glossectomy histopathological report after a previous excisional biopsy outside of MSK.

### Toxicity assessment

No clinically significant alterations in vital signs were observed in patients after PARPi-FL administration (see Supplementary Table 1). Temperature, blood pressure, respiratory rate and pulse remained similar or underwent changes within the physiological range pre and post swish. Patients who presented with hypertension before PARPi-FL administration continued to show elevated blood pressure after the procedure. The patient’s oral mucosa was checked by the responsible surgeon before PARPi-FL administration, immediately after imaging and 2 - 3 days later. No patients presented any signs of local irritation or reported signs of local toxicity when examined or actively questioned. Clinical blood sample analysis, encompassing complete blood count and differential, routine clinical chemistry and a metabolic panel revealed no pathological alterations related to PARPi-FL topical application.

### Analysis of PARPi-FL in Blood

No PARPi-FL was detected in the blood sample taken after the PARPi-FL swish via LCMS analysis in all patients. We determined the detection limit of our LCMS approach at 0.001 ng/μL of PARPi-FL and potential corresponding metabolites. Complete absorption of topically applied PARPi-FL into the bloodstream would correspond to 0.002 ng/μL at the highest dose level (1000 nM, assuming an average blood volume of 5 L), suggesting that measurable amounts of PARPi-FL after topical application are unlikely to enter the blood stream (Supplementary Fig. 2).

### Image acquisition optimization

Prior to patient imaging, we conducted tests with PARPi-FL containing agarose phantoms (0 - 5 μM PARPi-FL) to identify suitable settings for patient imaging (Supplementary Fig. 3). We aimed at optimizing the imaging settings towards sensitivity in the relevant concentration range of 100 - 1000 nM. As expected, the recorded signal intensity was lower at longer distance from the object (23 mm vs. 5 mm) and shorter exposure times (12 ms < 30 ms < 50 ms). To achieve a signal to background ratio of 3, which we considered adequate contrast to differentiate tumor from background tissue *in vivo*, at 23 mm from the object the PARPi-FL concentration needed to be 0.6 μM at 83 ms and 0.8 μM at 50 ms exposure time. Hence, this distance was considered incompatible to achieve sufficient image contrast at relevant concentrations. At 5 mm distance, the values decreased to 0.3 μM at 50 ms and 0.5 μM at 30 ms to achieve a signal to background ratio of 3 (Supplementary Fig. 3A). Since it was difficult to steadily hold and focus the endoscope at a 5 mm distance in patients, we chose a 10 mm distance from object for the clinical study and a 30 ms exposure time, allowing us to conduct lag-free real-time *in vivo* imaging.

**Figure 3.**
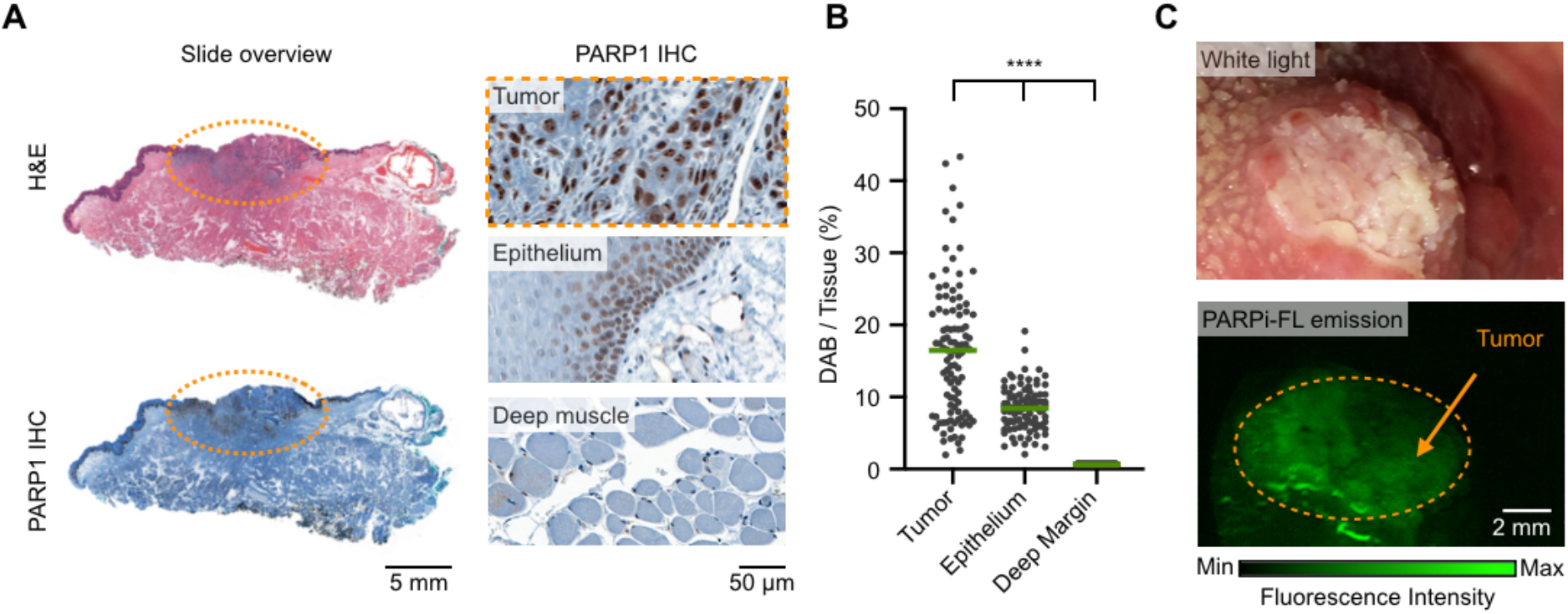
PARP1 expression in patients and PARPi-FL mediated contrast. **A**, Representative PARP1 immunohistochemistry images and H&E image from a biopsy of a patient (slide overview), showing an area of OSCC (orange circle). **B**, Quantification of PARP1 expression in immunohistochemistry samples (n = 8 patients) was performed and compared between tumor, epithelium, and deep margin. **C**, Phase I PARPi-FL imaging of the patient shown in panel A.

### PARP1 expression in patient tissue

Tissue samples from 8 patients were available for PARP1 expression analysis via IHC. Two of these specimens were excluded, since they had no tumor left in the paraffin block after standard-of-care histopathological assessment. PARP1 IHC staining was analyzed and quantified in tumor, mucosal epithelium and deep margin (i.e. healthy muscle tissue) based on the pathological assessment (**Fig. 3A**). PARP1 expression in tumor,16.35 ± 9.18% PARP1 area (DAB)/total tissue area (tissue), was significantly higher than in the normal mucosal epithelium (8.41 ± 2.91% DAB/tissue) and in the deep muscular margin (3.12 ± 1.79% DAB/tissue) (**Fig. 3B**; p < 0.0001 using linear regression analysis which considered the repeated measurements from the same patients, Supplementary Tables 2 and 3). On average, the % DAB/tissue was 45% lower in normal squamous epithelium compared to tumor and 81% lower in normal deep muscle compared to tumor.

### PARPi-FL imaging protocol

Groups of 3 patients were studied with increasing concentrations of PARPi-FL (100 nM, 250 nM, 500 nM, and 1000 nM). Patient 5 (250 nM) was analyzed separately, since the final pathology report did not identify any residual viable tumor. **Fig. 3C** shows an example of tumor enhancement with PARPi-FL at 250 nM. Signal quantification was conducted by placing ROIs on tumor and adjacent non-tumor tissue on still images of the three imaging time points “pre PARPi-FL”, “PARPi-FL pre-wash” and “PARPi-FL post-wash” and analysis of their **t**umor to normal **m**ucosal **r**atio (TMR). **Fig. 4A** shows an example of the signal increase in the tumor area after PARPi-FL application in a patient imaged at 1000 nM.

**Figure 4.**
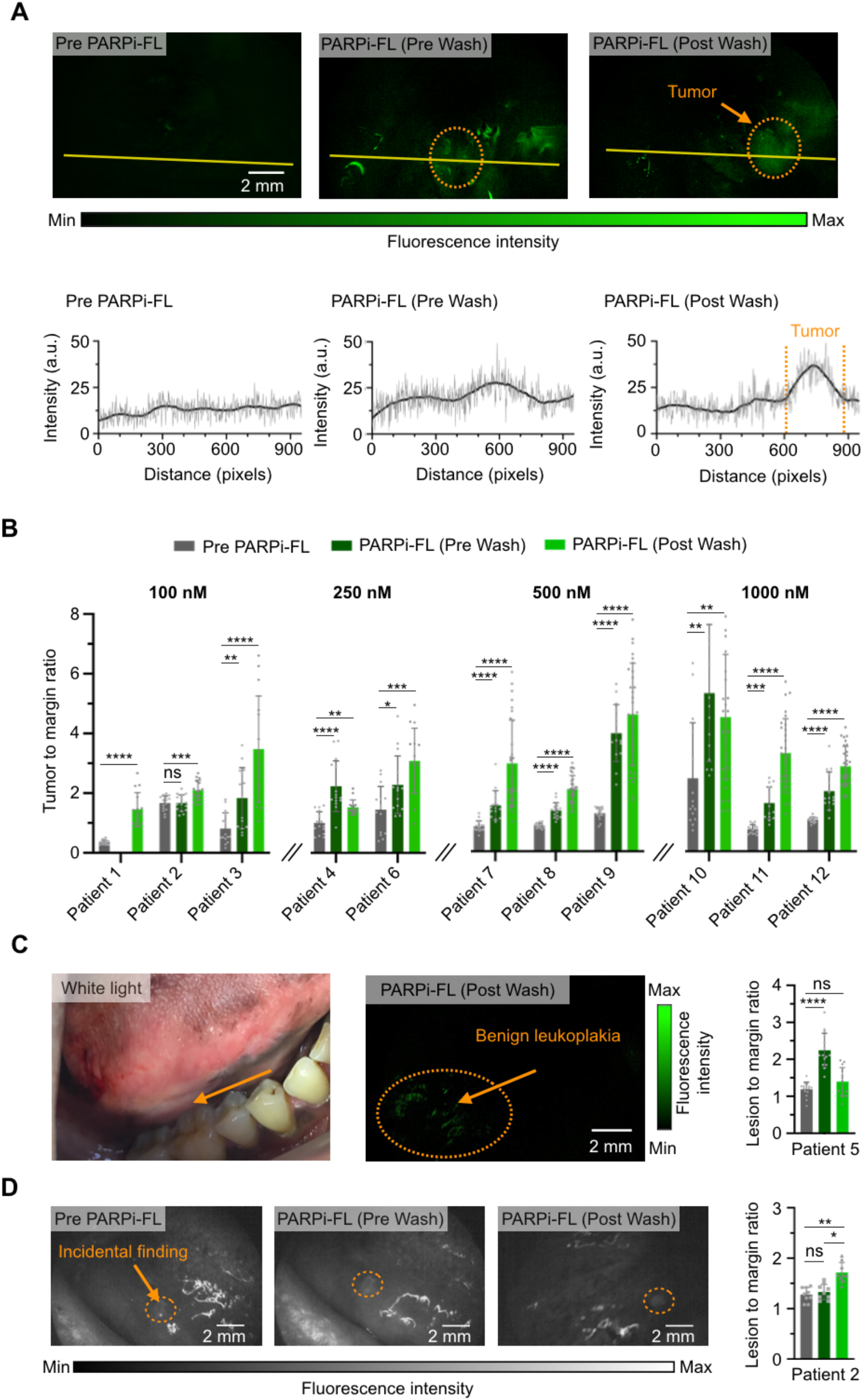
Intra-patient analysis after PARPi-FL imaging. **A**, Tumor area of a patient was imaged using a Quest Spectrum imaging device with an endoscopic camera pre-PARPi-FL, after gargling 1000 nM PARPi-FL solution (PARPi-FL, pre wash), and after gargling 500 nM PARPi-FL solution and a clearing solution of 30% PEG300 in PBS (PARPi-FL, post wash). The 2-D fluorescence intensity histogram illustrates the signal enhancement in the tumor area. **B**, Tumor-to-margin ratios of fluorescence imaging for each patient with histologically-proven OSCC (n = 11) pre PARPi-FL, after PARPi-FL (pre-wash) and after PARPi-FL (post-wash). Statistical significance was determined using the Wilcoxon matched-pairs signed rank test. \**P* < 0.05, \*\**P* < 0.01, \*\*\**P* < 0.001, \*\*\*\**P* < 0.0001. Data are presented as mean ± SEM. **C**, PARPi-FL post-wash image and quantification of patient 5. Histopathology found no OSCC. **D**, PARPi-FL imaging and quantification of the tumor-contralateral side of the tongue in patient 2. An intraoperative biopsy of the lesion was performed and confirmed a second primary 2 mm squamous cell carcinoma.

### Intra-patient imaging analysis

We compared TMR values pre PARPi-FL, PARPi-FL pre wash and PARPi-FL post-wash in each patient. A significant increase in TMR from pre PARPi-FL to PARPi-FL pre-wash (9/11 patients), and PARPi-FL post-wash (11/11 patients) was observed. The individual increase in TMR is displayed in **Fig. 4B**. The values on signal intensity and p-values between imaging time points for each patient can be found in Supplementary Table 4.

Our observations suggest that the clearing step is important in obtaining high contrast images. Without the clearing step, 18% (2/11) of the lesions would have been missed. We believe that this fact is due to the accumulation of unbound PARPi-FL around the tongue’s filiform papillae. The clearing step, associated with the saliva outflow, plays an important role in clearing the unbound compound before signal quantification is performed.

### Patient with no residual tumor in the surgical resection specimen

Patient 5 (250 nM), had a previous excisional biopsy of a tongue SCC with compromised margins at an outside institution. The patient presented with a flat residual lesion at the site of the previous resection that was assumed to be a persistent tumor. The patient had a pre-PARPi-FL lesion to margin ratio (LMR) of 1.20. The contrast increased after PARPi-FL administration (LMR: 2.24) but the contrast decreased after the clearing step (LMR: 1.53) (**Fig. 4C**).

### Patient with contralateral, incidentally identified lesion

Patient 2 had a large lesion on the lateral border of the tongue that was examined with PARPi-FL at the 100 nM dose level. In the process of imaging the contralateral side of the tongue in this patient as the imaging control, we noticed an area of increased PARPi-FL uptake (**Fig. 4D**). This area did not present any clinical evidence of macroscopic tumor in the pre-surgical setting. This area had TMRs of 1.28 (pre PARPi-FL), 1.33 (PARPi-FL pre-wash) and 1.71 (PARPi-FL post-wash). Post-wash, the TMR was significantly higher (p = 0.03) than pre PARPi-FL. Based on the pre-operative PARPi-FL finding and surgeon’s scrutiny after the patient was under anesthesia, intraoperative biopsy of this lesion was performed. Histopathologic examination confirmed a second primary squamous cell carcinoma at this location measuring 2 mm in the largest diameter.

### Inter-patient imaging analysis

We also conducted an analysis between the different PARPi-FL dose groups and observed a consistent increase in TMR in PARPi-FL pre-wash and post-wash images with increasing PARPi-FL dose levels (**Fig. 5A**). The mean predicted TMRs PARPi-FL post wash were 2.1 (100 nM), 2.1 (250 nM) and 2.8 (500 nM). At the highest dose level of 1000 nM PARPi-FL post wash yielded an average TMR of 3.3, a contrast value that is suitable to clearly distinguish tumor from normal tissue. Considering all time points, when accounting for the repeated measurements per patients (with a random intercept), the TMR was on average 39% higher at 250 nM compared to 100 nM, 62% higher at 500 nM than 100 nM dose, and 92% higher at 1000 nM as compared to 100 nM. These results indicated that 1000 nM was the most suitable concentration for PARPi-FL imaging. For descriptive data on the linear regression analysis, the average values on TMR, and the p-values please see Supplementary Table 5.

**Figure 5.**
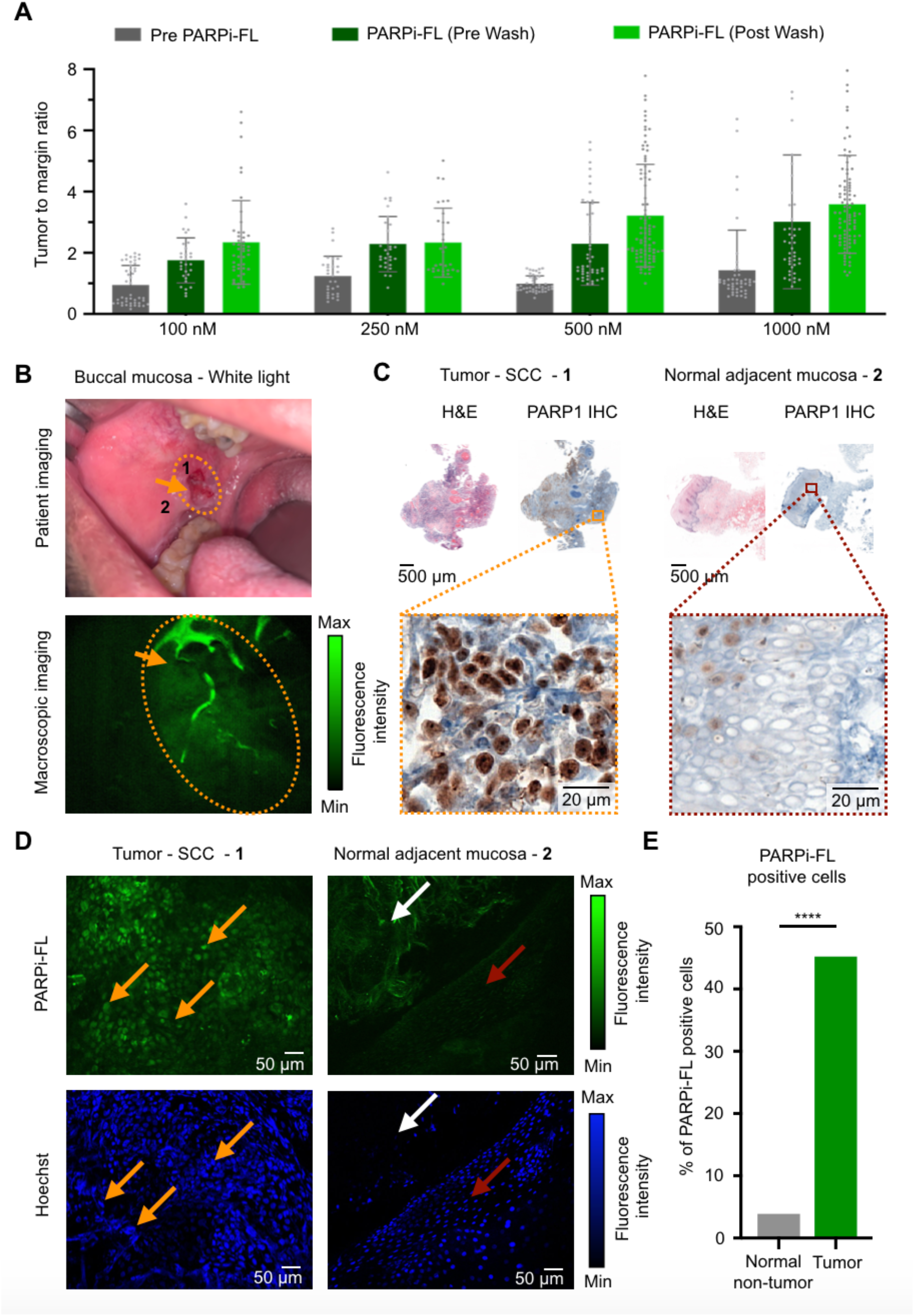
Inter-patient analysis and subcellular specificity of PARPi-FL. **A**, Tumor-to-margin ratio of fluorescence imaging for dose groups (100 nM, 250 nM, 500 nM, and 1000 nM). Ratios were calculated from 5 ROIs per FOV and 3 FOVs of each patient. Displayed are individual data points, means and SEM. **B**, PARPi-FL macroscopic imaging of a patient sample from phase II (PARPi-FL post-wash, 1000 nM), which was selected for microscopic confirmation of PARPi-FL specificity. 1 = area of tumor where the biopsy was taken from, 2 = area where the biopsy of the free-of-disease margin was taken from. **C**, H&E and PARP1 IHC (overview and zoom-in) of the tumor and margin biopsies from patient in panel B, demonstrating higher PARP1 expression in the tumor area compared to normal adjacent mucosa. **D**, Microscopic analysis of the tumor and margin biopsies to evaluate the PARPi-FL accumulation following topical 1 min swish & spit application. Fresh tissues underwent nuclear counterstaining with Hoechst 33342 *ex vivo* prior to microscopy. Orange arrows point to the nuclei of tumor cells, red arrows point to nuclei of cells in a normal (benign) basal layer, and white arrows point to the interstitial layer (collagen autofluorescence). **E**, Quantification of PARPi-FL fluorescence signal inside the nuclei of cells. Cell nuclei were identified using Hoechst 33342.

### Microscopic imaging of PARPi-FL accumulation

To demonstrate that the macroscopically detected increase in fluorescence signal after PARPi-FL application was based on its specific accumulation in cell nuclei, we analyzed a tumor and free-of disease normal tissue sample originating from a patient on phase II. The patient harbored a right buccal mucosal OSCC and was imaged after 1000 nM PARPi-FL and clearing with 30% PEG 300 in water (**Fig. 5B**). IHC corroborated the difference in PARP1 expression within the tumor and the free-of-disease normal tissue in this patient (**Fig 5C**). Confocal microscopy images clearly demonstrated that PARPi-FL accumulated exclusively in tumor cell nuclei, while no nuclear signal was observed in the non-tumor tissue (**Fig. 5D**). Quantification was carried out by verifying the presence of PARPi-FL fluorescence signal inside the nucleus of cells. In the tumor tissue 1061 nuclei were quantified and 45.14% of cells showed PARPi-FL uptake. In the normal tissue 412 nuclei were analyzed) and only 3.89% of cells had PARPi-FL uptake. This difference was statistically significant (p < 0.0001), corroborating the specificity of PARPi-FL for tumor cells. (**Fig. 5E**).

## Discussion

Fluorescence-guided imaging using molecularly targeted agents to delineate tumors for diagnosis and to guide surgical resection has started to enter the clinical research space in recent years (30). These include studies on ovarian, colorectal and breast cancer (31). While only late stage clinical studies will be able to prove the added clinical value in terms of survival benefit of these newer modalities compared to standard diagnosis and surgical resection, early phase clinical studies are crucial to show feasibility of lesion detection, dose-finding, and provide exploratory data on sensitivity and specificity. In this regard, tumor-to-background ratios, imaging contrast and signal specificity are important readouts in phase I and II studies. In this study, we report the clinical data from a phase I dose-escalation study of PARPi-FL, where we focus on safety and imaging contrast after topical application in oral cancer patients.

Several other fluorescent agents (such as γ-glutamyl hydroxymethyl rhodamine green (32), indocyanine green (33), urokinase-like plasminogen activator receptor (34) cetuximab/panitumumab-IRDye800 (35,36)) have been previously investigated for oral cancer delineation and surgical margin assessment. Other, non-targeted diagnostic aids have also been suggested for oral cancer detection, such as light-based detection systems (11,14,37) and non-fluorescent dyes (such as Lugol’s iodine (38) and toluidine blue (39)). Nevertheless, their clinical value remains unclear and their adoption into clinical use has been low (9,40). We have recently identified PARP1 as a biomarker for OSCC, based on its overexpression in patient tissue (20,25,27). Our current approach is therefore focused on leveraging this finding in development of a diagnostic aid for early detection of OSCC.

We have established the high specificity of the fluorescent PARP1-targeted imaging agent PARPi-FL in mouse models of oral cancer (41-43) and in human tissues (25) for its ability to delineate malignant from non-malignant tissue. In this current study, we have used PARPi-FL as a topical swish & spit solution, which patients gargled for one minute before imaging. The highest PARPi-FL dose in our study, 1000 nM, is equivalent to 15 nmol, which qualifies as a microdose under FDA guidelines.

Our phase I study in 12 patients revealed that PARPi-FL topical application is safe, with no adverse events or signs of local or systemic toxicity in any patient. Regarding the dose-finding objective of the study, we found that the highest dose of PARPi-FL, 1000 nM, led to the most pronounced differential in contrast between tumor and surrounding non-tumor tissue, consistently crossing the threshold of a TMR (=contrast) of > 3. TMRs of 3 - 4, as achieved in this study, provide a level of optical contrast that is reliably apparent to the human observer, and which may not need further computational or statistical analysis to distinguish tumor from non-tumor tissue (44).

Here, we first optimized the instrument settings towards highest sensitivity on agarose phantoms and then applied the selected imaging parameters throughout the study to ensure comparability between patients and dose levels. In addition, our results indicate that a 1 min clearing step after the PARPi-FL application was important to maximize contrast and reduce false positive as well as false negative results. In fact, TMR values were consistently higher post-wash, particularly at the two highest dose-levels.

Furthermore, several incidental findings in our study suggest that our approach is specific and sensitive, although a thorough evaluation of these parameters was outside the scope of a phase I study. We observed that the only patient who did not show a significant increase in TMR between the pre-PARPi-FL and PARPi-FL post-wash imaging in fact did not have any residual tumor in the surgical specimen of definitive glossectomy after a pre-surgical excision biopsy. In addition, we identified a previously unrecognized cancer on the contralateral side of the tongue based on PARPi-FL contrast, which then directed the surgeon to examine that area and perform a biopsy during surgery which led to early diagnosis of a clinically inapparent tumor. Although more studies are necessary to evaluate PARPi-FL uptake in benign and dysplastic lesions, our results to date are encouraging. Lastly, we were able to confirm on a cellular level that PARPi-FL accumulated in tumor cell nuclei following the 1 min swish & spit protocol, strongly supporting the specificity of the macroscopic TMR increase that we have observed on optical imaging.

A central aspect of PARPi-FL-based oral cancer detection is the fact that the lesions of interest are located at or close to the tissue surface. Importantly, our goal is to determine whether a visible lesion is of malignant or non-malignant nature and whether it needs to be biopsied or treated.

Therefore, the green emission spectrum of PARPi-FL, which prohibits deep tissue penetration but provides excellent brightness, is eminently suitable for clinical translation. We previously discussed the design constraints of fluorescent PARP inhibitors in-depth, outlining specifically why near-infrared labels have not been successfully implemented in PARP1 imaging approaches (26,45,46). We believe that the expression of PARP1, coupled with the pharmacokinetic profile and potential clinical benefit of diagnostic imaging with PARPi-FL far outweigh the trade-offs associated with its wavelengths (25). This is supported by other clinical studies that involve green fluorescent dyes, including the first-in-human study of a molecularly targeted optical imaging agent (47) and the recent introduction of a green-fluorescent confocal microscope for brain tumor imaging (48). In the present study, we used a customized imaging system equipped with a FITC LED and bandpass filters. The broader autofluorescence peak width allowed us to separate specific (PARPi-FL) and non-specific (autofluorescence) emissions in a post-processing step. For future directions, additional refinement of the imaging software will be necessary to allow specific PARPi-FL imaging in real time. Importantly, successful transfer of this technology to low-resource health care settings in under-developed or developing geographic areas that have a high prevalence of oral lesions requires a modification of the imaging system such that a cost-effective hand-held device can be used even by trained non-medical personnel to facilitate early detection of OSCC within well-designed screening programs.

Topical application of PARPi-FL is a rapid, specific, and safe technique for *in vivo* detection of OSCC. No toxicities were reported or observed in any of the study participants. The specificity of the technique was supported by our finding that topically applied PARPi-FL accumulated preferentially within nuclei of tumor cells, corroborating our imaging findings at the subcellular level. We have identified 1000 nM as the optimal dose for the Phase II of the clinical study, yielding the highest tumor-to-normal ratio. In conclusion, PARPi-FL imaging is a safe, rapid, *in vivo*, non-invasive (no contrast agent injection) technique that has the potential for universal implementation for evaluation of suspicious lesions in the oral cavity and early detection of OSCC.

## Data Availability

All data from this study are presented in the main manuscript and Supplementary information. De-identified raw data can be made available upon reasonable request.

## Author Contributions

P.D.S.F., S.K., C.B; S.P., and T.R. conceived the study and designed the experiments. P.D.S.F, S.K., C.B, N.G., D.K.Z., D.A., C.V., R.G.G., H.S., I.G., S.P., and T.R. carried out the experiments and collected the data. C.B., S.R. and T.R. produced GMP PARPi-FL. P.D.S.F., C.B., R.A.G., S.P., and T.R. analyzed the data. P.D.S.F., S.K., C.B., N.G., W.A.W.; S.P., and T.R. wrote IRB protocols. P.D.S.F., C.B., A.M., S.P., and T.R. conducted statistical analysis of the data. P.D.S.F., S.K., C.B., and T.R. primarily wrote the manuscript. All authors carefully read and edited the manuscript.

## Acknowledgements

The authors gratefully acknowledge the support of the Memorial Sloan Kettering Cancer Radiochemistry & Molecular Imaging Probes Core, the Molecular Cytology Core and the Memorial Sloan Kettering Center for Molecular Imaging & Nanotechnology. We especially thank Eric Chan for the help with the Image J plugins for PARP1 and PARPi-FL quantifications. We also thank Garon Scott for proofreading and Terry Helms for creating illustrations. We also thank the operating room nurses and the Head and Neck surgical faculty and clinical staff at MSK, especially Violeta Dokic, R.N. for the help with the patients.

