## Supplementary Information for "A Phase I Study of a PARP1-targeted Topical Fluorophore for the Detection of Oral Cancer"

1275 York Avenue

New York, NY, 10065

1-646-888-3461

SUPPLEMENTARY FIGURES AND TABLES:

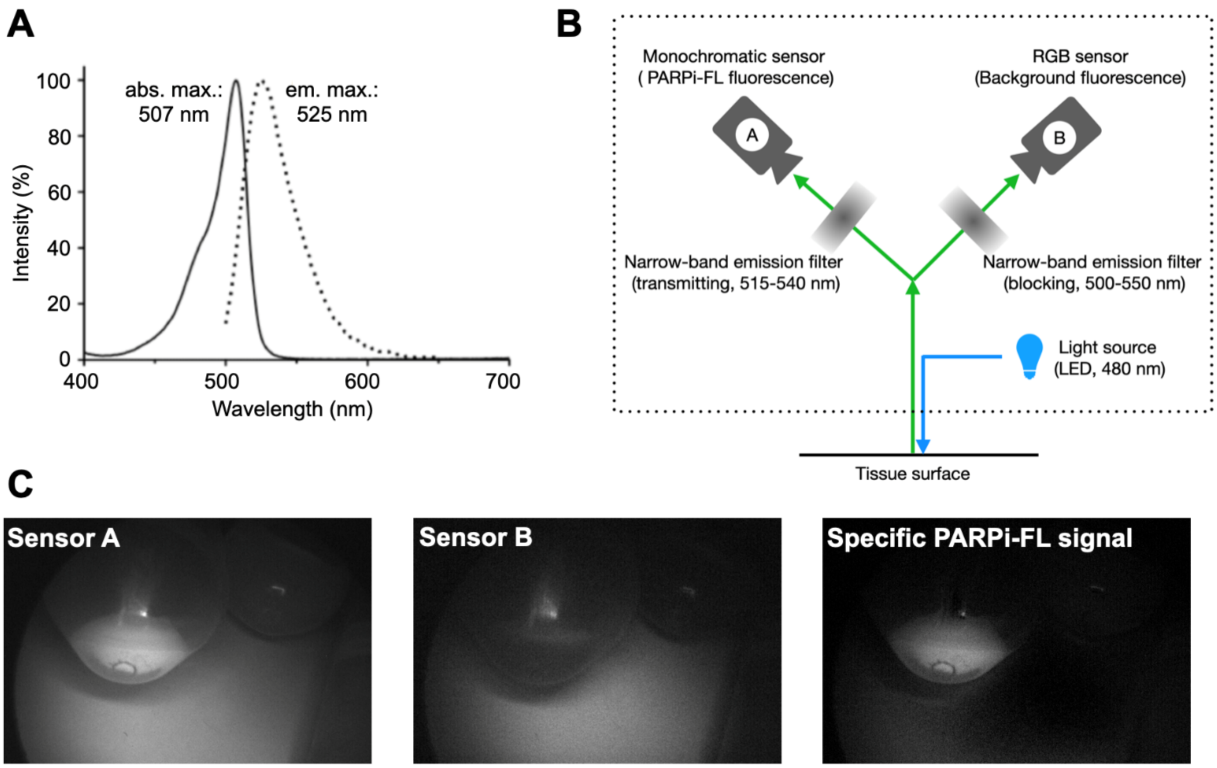


**Supplementary Figure S1.**

Fluorescence imaging system. **A,** The specific signal (PARPi-FL) can be separated from the non-specific (autofluorescence) emissions due to the broader autofluorescence peak width. **B,** Schematic representation of PARPi-FL signal acquisition. **C,** The obtained images were post-processed to separate specific (PARPi-FL, sensor A) and non-specific (autofluorescence, sensor B) emissions based on different peak widths of the emission spectra.

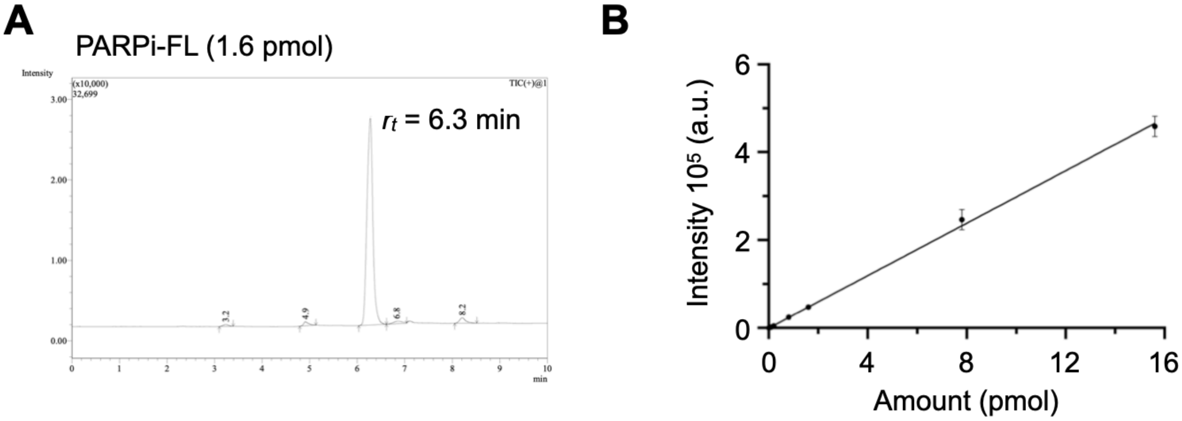


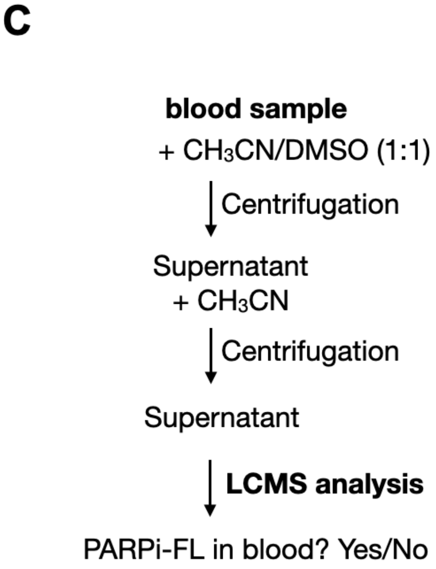


D

| Blood sample | LCMS signal at 6.3 min |
|---|---|
| Patient 1 | Not detected |
| Patient 2 | Not detected |
| Patient 3 | Not detected |
| Patient 4 | Not detected |
| Patient 5 | Not detected |
| Patient 6 | Not detected |
| Patient 7 | Not detected |
| Patient 8 | Not detected |
| Patient 9 | Not detected |
| Patient 10 | Not detected |
| Patient 11 | Not detected |
| Patient 12 | Not detected |

**Supplementary Figure S2.**

Blood sample analysis after topical administration of PARPi-FL **A,** LCMS chromatogram of PARPi-FL (1.6 pmol). **B,** LC/MS analysis of varying amounts of PARPi-FL. **C,** Workflow for analyzing research blood samples from clinical phase I trial after topical administration of PARPi-FL. **D,** Results for each blood sample of patients 1 through 12, yielding no measurable amounts of PARPi-FL in the plasma.

**Supplementary Table S1.**

PARPi-FL Toxicity assessment on Vital signs. Conc. = Concentration, IPM = incursions per minute, BPM = beats per minute, MBP = mean blood pressure, RR = respiratory rate, Temp = frontal head temperature.

**Phase I - Vitals pre and post swish of PARPi-FL**

| Patient identification | | | | Vitals pre-swish | | | | Vitals post-swish | | | | ToxicityToxicity | |
|---|---|---|---|---|---|---|---|---|---|---|---|---|---|
| Case | (nM) | Height (cm) | Weight (Kg) | Temp (°C) | MBP (mmHg) | RR (IPM) | Pulse (BPM) | Temp (°C) | MBP (mmHg) | RR (IPM) | Pulse (BPM) | D 0 | D 2-4 |
| 1 | 100 | 1.6 | 67.7 | 37 | 159 | 95%* | 77 | 36.8 | 153 | 98%* | 75 | no | no |
| 2 | 100 | 1.8 | 95.3 | 36.6 | 95 | 18 | 70 | 36.2 | 96 | 18 | 75 | no | no |
| 3 | 100 | 1.55 | 90.7 | 36.9 | 97 | 20 | 92 | 36.4 | 99 | 24 | 87 | no | no |
| 4 | 250 | 1.65 | 107 | 36.7 | 123 | 20 | 66 | 36.5 | 100 | 20 | 61 | no | no |
| 5 | 250 | 1.7 | 67.1 | 36.6 | 112 | 20 | 87 | 36.6 | 86 | 20 | 69 | no | no |
| 6 | 250 | 1.67 | 56.2 | 36.7 | 77 | 98%* | 63 | 36.7 | 77 | 99%* | 61 | no | no |
| 7 | 500 | 1.7 | 69.8 | 36.9 | 119 | 20 | 81 | 36.9 | 125 | 20 | 74 | no | no |
| 8 | 500 | 1.57 | 86.2 | 37 | 114 | 20 | 77 | 36.8 | 93 | 20 | 71 | no | no |
| 9 | 500 | 1.77 | 47.4 | 37.1 | 90 | 20 | 69 | 37.1 | 118 | 20 | 74 | no | no |
| 10 | 1000 | 1.7 | 85.2 | 36.8 | 107 | 18 | 86 | 36.8 | 112 | 20 | 93 | no | no |
| 11 | 1000 | 1.8 | 82.5 | 37 | 95 | 18 | 77 | 37 | 101 | 18 | 76 | no | no |
| 12 | 1000 | 1.78 | 83.9 | 36.9 | 105 | 16 | 66 | 36.6 | 115 | 18 | 58 | no | no |

*Note that 2 patients did not have their respiratory rate taken but the SaO2. First case SaO2 went from 95% pre PARPi-FL administration to 98% post-administration. The second case it went from 98% to 99%. In both cases values were within the normal range.

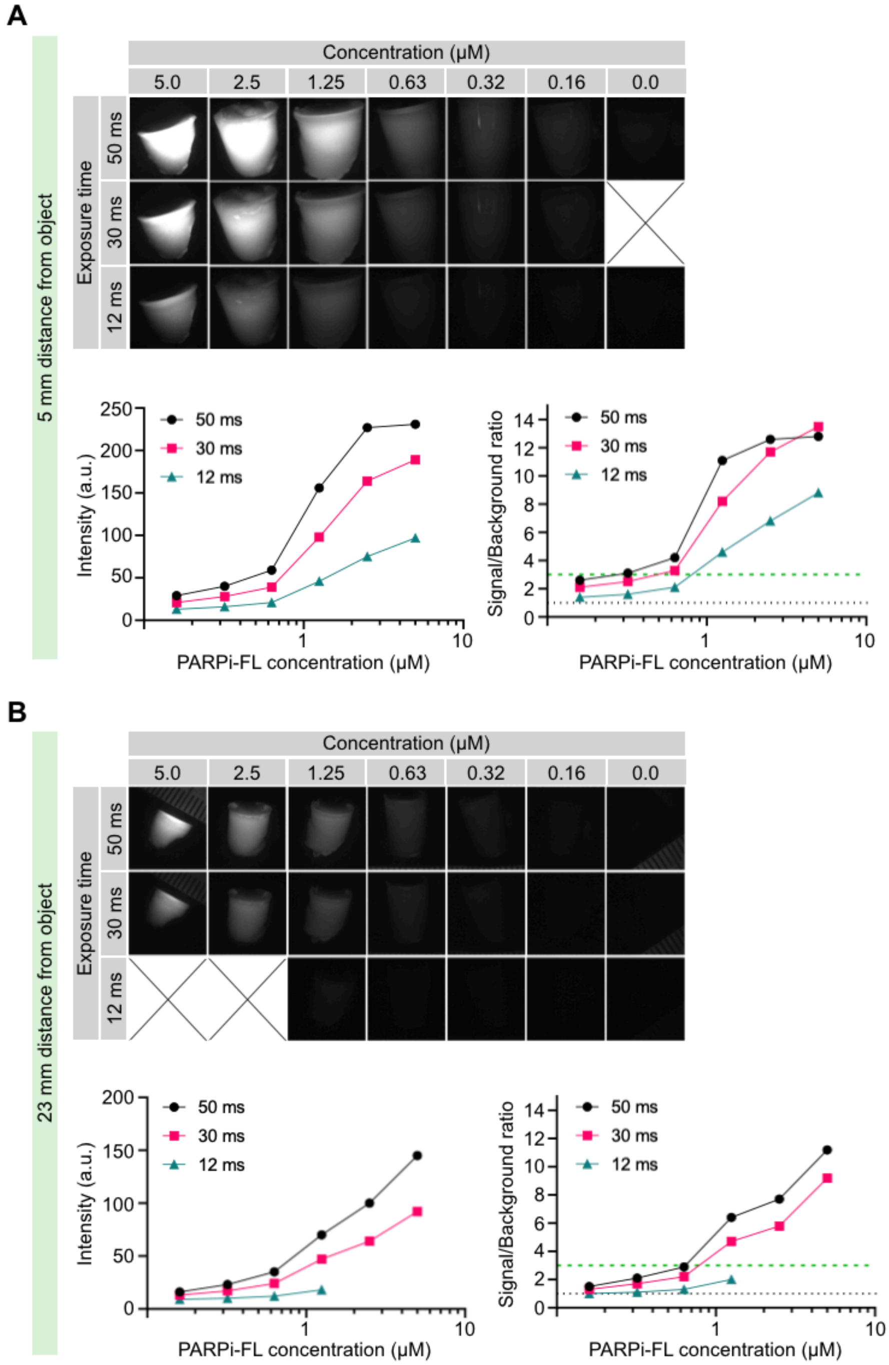


**Supplementary Figure S3.**

Evaluation of sensitivity of the imaging system using PARPi-FL containing agarose phantoms (0-5000 nM), varied exposure time (12 ms, 30 ms, and 50 ms), and distance from object (5 mm vs. 23 mm). **A,** Fluorescence images and quantification using a distance of 5 mm. **B,** Fluorescence images and quantification using a distance of 23 mm.

**Supplementary Table S2.**

Quantification of PARP1 expression on IHC slides.

| Tissue | Measures (n) | Patient (n) | Mean | SD | Median | Min | Max |
|---|---|---|---|---|---|---|---|
| Epithelium | 105 | 8 | 8.41 | 2.91 | 8.46 | 2.04 | 19.16 |
| Margin | 212 | 8 | 3.12 | 1.79 | 2.89 | 0.3 | 12.71 |
| Tumor | 102 | 6 | 16.35 | 9.18 | 16.5 | 1.98 | 43.34 |

**Supplementary Table S3.**

Linear regression to account for the repeated measurements per patient.

| Tissue | Predicted average value | Coefficient | 95% CI | p value |
|---|---|---|---|---|
| Tumor | 14 | 1.00* | | < 0.0001 |
| Epithelium | 7.8 | 0.55 | 0.48, 0.64 | |
| Margin | 2.7 | 0.19 | 0.17, 0.21 | |

*Reference value

**Supplementary Table S4.**

Quantification of PARPi-FL signal at different concentrations.

| PARPi-FL concentration | Patient # | Average signal (a.u.) ± SD | | | Signal Increase (times) | | p-value | |
|---|---|---|---|---|---|---|---|---|
| | | Pre-PARPi-FL | Post-PARPi-FL Pre-Wash | Post-PARPi-FL Post-wash | Post-PARPi-FL Pre-Wash | Post-PARPi-FL Post-wash | Post-PARPi-FL Pre-Wash | Post-PARPi-FL Post-wash |
| *100 nM* | 1 | 0.36 ± 0.07 | N/A | 1.46 ± 0.56 | N/A | 4.10 | N/A | < 0.0001 |
| | 2 | 1.67 ± 0.25 | 1.67 ± 0.27 | 2.1 ± 0.33 | 1.00 | 1.26 | 0.9341 | 0.0009 |
| | 3 | 0.82 ± 0.53 | 1.84 ± 1.02 | 3.48 ± 1.78 | 2.24 | 4.24 | 0.0012 | < 0.0001 |
| *250 nM* | 4 | 1.02 ± 0.38 | 2.26 ± 0.86 | 1.55 ± 0.25 | 2.24 | 1.52 | < 0.0001 | 0.0012 |
| | 5* | 1.19 ± 0.19 | 2.24 ± 0.46 | 1.53 ± 0.48 | 1.88 | 1.28 | < 0.0001 | 0.0554 |
| | 6 | 1.47 ± 0.79 | 2.31 ± 0.98 | 3.13 ± 1.10 | 1.57 | 2.13 | 0.0151 | 0.0001 |
| *500 nM* | 7 | 0.83 ± 0.19 | 1.54 ± 0.49 | 2.96 ± 1.45 | 1.85 | 3.55 | < 0.0001 | < 0.0001 |
| | 8 | 0.86 ± 0.08 | 1.37 ± 0.26 | 2.09 ± 0.45 | 1.59 | 2.44 | < 0.0001 | < 0.0001 |
| | 9 | 1.28 ± 0.21 | 3.97 ± 0.95 | 4.6 ± 1.72 | 3.11 | 3.60 | < 0.0001 | < 0.0001 |
| *1000 nM* | 10 | 2.48 ± 1.88 | 5.36 ± 2.32 | 4.55 ± 2.12 | 2.16 | 1.84 | 0.002 | 0.0015 |
| | 11 | 0.75 ± 0.15 | 1.64 ± 0.54 | 3.33 ± 1.19 | 2.18 | 4.43 | 0.0001 | < 0.0001 |
| | 12 | 1.07 ± 0.09 | 2.06 ± 0.64 | 2.89 ± 0.67 | 1.94 | 2.71 | < 0.0001 | < 0.0001 |

*Patient did not have viable tumor in the final anatomopathological report

**Supplementary table S5.**

Change of TMR values in within dose cohorts.

| Time | Average value | Coefficient | 95% CI | p-value |
|---|---|---|---|---|
| ***100 nM*** | | | | |
| Pre PARPi-FL | 0.7 | 1.00 (Ref.) | | <0.0001 |
| Post PARPi-FL (Pre wash) | 1.2 | 1.67 | 1.32, 2.14 | |
| Post PARPi-FL (Post wash) | 2.1 | 2.83 | 2.31, 3.47 | |
| ***250 nM*** | | | | |
| Pre PARPi-FL | 1.1 | 1.00 (Ref.) | | <0.0001 |
| Post PARPi-FL (Pre wash) | 2.1 | 1.96 | 1.57, 2.45 | |
| Post PARPi-FL (Post wash) | 2.1 | 1.94 | 1.56, 2.43 | |
| ***500 nM*** | | | | |
| Pre PARPi-FL | 1 | 1.00 (Ref.) | | <0.0001 |
| Post PARPi-FL (Pre wash) | 2 | 2.06 | 1.80, 2.37 | |
| Post PARPi-FL (Post wash) | 2.8 | 2.97 | 2.63, 3.34 | |
| ***1000 nM*** | | | | |
| Pre PARPi-FL | 1.2 | 1.00 (Ref.) | | <0.0001 |
| Post PARPi-FL (Pre wash) | 2.5 | 2.14 | 1.79, 2.55 | |
| Post PARPi-FL (Post wash) | 3.3 | 2.85 | 2.44, 3.32 | |
